# Germline cancer gene expression quantitative trait loci influence local and global tumor mutations

**DOI:** 10.1101/2022.08.23.22279002

**Authors:** Yuxi Liu, Alexander Gusev, Peter Kraft

**Author notes:** **Corresponding author:** Peter Kraft, PhD. Program in Genetic Epidemiology and Statistical Genetics, Harvard T.H. Chan School of Public Health, 655 Huntington Avenue, Boston, MA, 02115.

## Abstract

Somatic mutations drive cancer development and are relevant to patients’ response to treatment. Emerging evidence shows that variations in the somatic genome can be influenced by the germline genetic background. However, the mechanisms underlying these germline-somatic associations remain largely obscure. We hypothesized that germline variants can influence somatic mutations in a nearby cancer gene (“local impact”) or a set of recurrently mutated cancer genes across the genome (“global impact”) through their regulatory effect on gene expression. We integrated tumor targeted sequencing from 12,413 patients across 11 cancer types in the Dana-Farber Profile cohort with germline cancer gene expression quantitative trait loci (eQTL) from the Genotype-Tissue Expression Project. We identified variants that upregulate *ATM* expression which are also associated with a decreased risk of having somatic *ATM* mutations across 8 cancer types (*P* = 3.43 × 10^−5^). We also identified *GLI2, WRN*, and *CBFB* eQTL that are associated with global tumor mutational burden of cancer genes in ovarian cancer, glioma, and esophagogastric carcinoma, respectively (*P* < 3.45 × 10^−6^). An *EPHA5* eQTL was associated with the number of mutations in cancer genes specific to colorectal cancer, and eQTL associated with expression of *APC, WRN, GLI1, FANCA*, and *TP53* were associated with mutations in genes specific to endometrial cancer (*P* < 1.73 × 10^−5^). Our findings provide evidence for the germline-somatic associations mediated through expression of specific cancer genes and open new avenues for research on the underlying biological processes, especially those related to immunotherapy responses.

## Introduction

Cancer is a genetic disease driven by somatic events occurring in the genome over time. Identifying genes carrying driver mutations (cancer driver genes) and elucidating their roles in the related signaling pathways have become primary goals in cancer genomic research because of the contribution of these genetic changes to abnormal and uncontrolled cell growth and transformation which drive the development of a malignant tumor (1-4). Many of these driver genomic alterations have been found to be clinically actionable drug or therapeutic targets for precision medicine. With the advancement of low-cost, high-throughput next-generation sequencing (NGS) technologies, genomic profiling of tumors using targeted NGS panels is becoming part of routine cancer care (5-8).

Different cancers are characterized by different patterns of somatic mutations (9,10). Even patients with the same cancer may have substantial heterogeneity in the overall tumor mutational burden (TMB), mutation patterns characterized by mutational signatures, or the cancer genes and oncogenic signaling pathways altered (4,11-14). These heterogeneities in the somatic mutational profile can lead to differential cancer progression, prognosis, and treatment response (15,16). A well-known example is the predictive association of TMB and response to immunotherapy (17). Mounting evidence suggests that somatic variations in tumors can have a germline genetic basis (12,18-23). This germline-somatic relationship has been established at different levels, from the impact of a single germline variant on somatic mutation rate of a cancer gene (e.g., rs25673 at 19p13.3 with *PTEN* alterations that involved in the mTOR signaling pathway) (20), to the associations between germline polygenic risk scores (PRS) and somatic mutational signatures (e.g., germline PRS of inflammatory bowel disease with APOBEC signatures in breast cancer) (23). Emerging evidence also shows an interactive effect of germline and somatic variations on clinical outcomes (24). However, the study of germline-somatic interactions is still at an early stage and the mechanisms responsible for these observed associations are still largely uncovered.

Germline variants may affect somatic mutations through gene expression (19,22,25). In the well-established example of the APOBEC mutational process, rs17000526-A allele in the *APOBEC3B* region is associated with higher expression of this gene, which contributes to somatic mutagenesis of APOBEC signatures in bladder tumor (19). Chen et al. systematically assessed the impacts of expression level of putative cancer-susceptibility genes on mutational signatures and TMB and identified a wide range of associations across six cancer types (25). Many underlying mechanisms may co-exist, but an intuitive and interpretable hypothesis would be that the germline cancer gene expression quantitative trait loci (eQTL) alter the propensity of acquiring somatic mutations in those specific genes or globally through their regulatory effect on gene expression. Although prior studies included gene expression information in the analysis of germline-somatic interactions, this is no systematic study focusing on both the local and global impact of eQTL on somatic mutations in cancer genes across multiple cancers. Many latent associations and mechanisms may thus have been missed.

Here, we performed a pan-cancer analysis of the germline genetic impacts on both the local and global tumor mutations, incorporating regulatory information of germline variants on gene expression. Specifically, we evaluated the associations between germline cancer gene eQTL and i) somatic mutation status of those cancer genes or any hotspot mutation in those genes, ii) tumor mutation counts (TMC) of all recurrently mutated cancer genes for a cancer type, and iii) TMB of all targeted cancer genes from the OncoPanel sequencing platform across 11 cancer types in the Dana-Farber Profile cohort. Clinical targeted sequencing cohorts are well suited for such germline-somatic analysis because the tumor sequencing specifically targets those actionable cancer drivers and the cancer patient population is usually large, unselected, and has extensive clinical data. Our results demonstrate evidence for germline-somatic associations that are potentially mediated through cancer gene expression and provide insights into the mechanisms of mutagenesis in somatic cells.

## Materials and Methods

### Study population

The Dana-Farber Profile, initiated in 2011, is a cohort study of unselected cancer patients who presented at the Dana-Farber Cancer Institute, Brigham and Women’s Hospital, or Boston Children’s Hospital, received genomic profiling and consented to participate. Tumor specimens, mainly formalin-fixed paraffin-embedded tissues, were retrieved from all consented patients for targeted sequencing. Comprehensive clinical and pathologic data were collected along with the genomic data (6,26). The study protocol was approved by the institutional review board (IRB) of Dana-Farber/Partners Cancer Care Office for the Protection of Research Subjects (11-104/17-000). Secondary analyses of previously collected data were approved by the Dana-Farber IRB (19-033/19-025).

### Tumor targeted sequencing

A workflow of the full data generating and processing pipeline is present in **Fig. 1**. All collected tumor samples were sequenced on OncoPanel, a targeted NGS platform designed for detecting somatic variations in a panel of actionable cancer genes. There are three versions of the panel targeting the exon and/or intron regions of 304, 326, and 447 genes, respectively; each patient in the cohort was sequenced on one of the panels (Supplementary Table S1). All targeted genes were previously identified oncogenes or tumor suppressor genes involved in cancer-related signaling pathways (27). Sequencing was performed using an Illumina HiSeq 2500 with 2×100 paired-end reads followed by somatic mutation calling using MuTect (for single-nucleotide variants) (28) and Indelocator (for indels; http://www.broadinstitute.org/cancer/cga/indelocator) from reads aligned to the targeted genome regions with > 50× reads (“On-target reads”). More details about the tumor sequencing pipeline can be found in prior studies (6,27).

**Figure 1.**
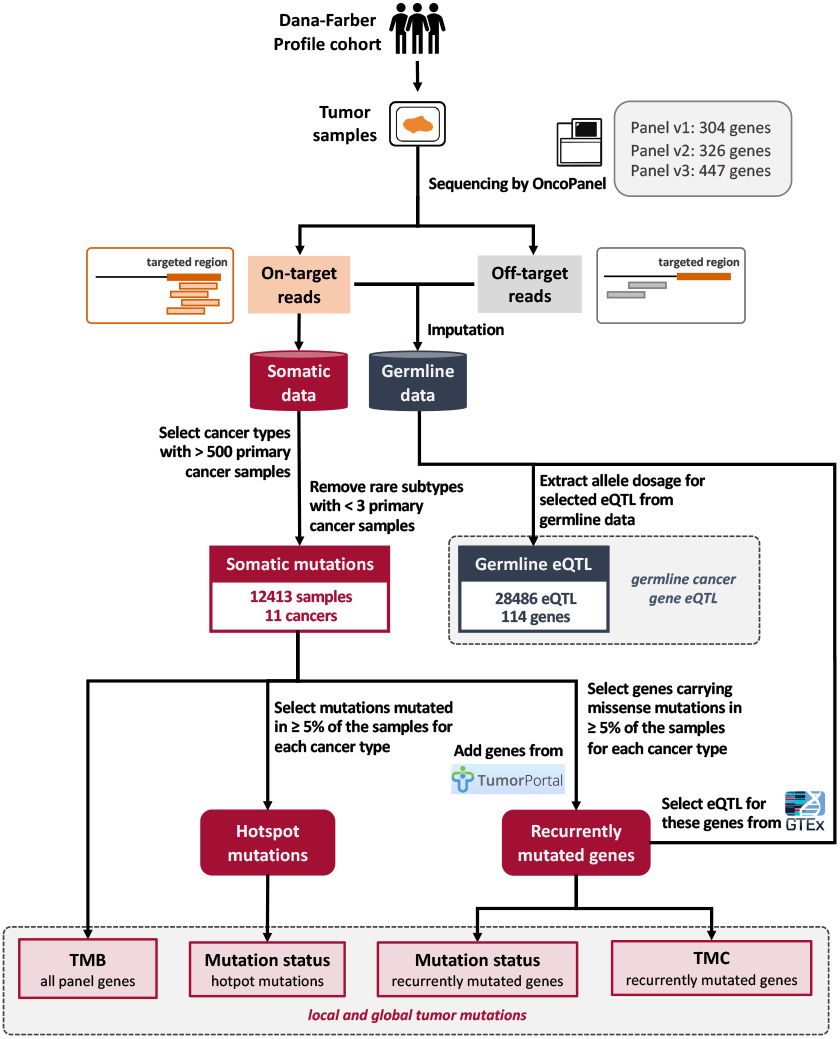
A workflow of the germline and somatic data generation pipeline in the Profile cohort. Tumor samples were collected from all consented patients in the Profile cohort, followed by targeted sequencing using OncoPanel. Somatic data were generated from on-target reads from the tumor sequences. Germline data were imputed using both the off-target and on-target reads generated from tumor sequencing. Four measures of the local and global tumor mutations: i) Mutation status of recurrently mutated cancer genes, ii) Mutation status of hotspot mutations, iii) TMC of recurrently mutated cancer genes, and iv) TMB of all panel genes were generated for all selected primary cancer samples across 11 cancer types from somatic data. Germline eQTL were identified from GTEx for all identified genes, followed by germline allele dosage extraction from the germline imputed data.

We collected somatic mutation data from the tumor sequences of 18,472 primary cancer samples spanning over 60 cancer types and subtypes. Some tumors exhibit microsatellite instability (MSI) with high mutational burden; the germline-somatic relationship for those hypermutable subtypes might be substantially different from the microsatellite stable (MSS) tumors. We thus further classified each sample as MSI or MSS using MSIDetect (29). Cancer types with > 500 samples were selected; for each selected cancer, we removed those rare subtypes with < 3 samples. The remaining 12,413 samples across 11 cancer types were included in the downstream analysis (Supplementary Table S2).

### Germline imputation from tumor sequences

Details of inferring common germline variants from the OncoPanel tumor sequencing data are described elsewhere (26) and briefly summarized here. Tumor targeted sequencing generated both high-coverage “on-target reads” aligned to the targeted regions and low-coverage “off-target reads” aligned to the rest of the genome (**Fig. 1**). Common germline variants with > 1% frequency in the European population were imputed from these tumor sequences (mainly relied on off-target reads) using linkage disequilibrium (LD) information with the 1000 Genomes Phase 3 release as the haplotype reference panel. Imputation accuracies from several algorithms designed for imputing germline variants from low coverage data were evaluated by comparing the imputed allele dosage to the gold standard germline data generated from genotyping array.

The STITCH algorithm (30) yielded the highest overall accuracy and the resulting imputed germline data were used for the downstream analysis. The imputed variants were subsequently restricted to an imputation INFO score of greater than 0.4, which produced a mean imputation correlation of 0.86 between tumor imputed and germline SNP array variants (26).

Genetic ancestry was inferred by projecting the imputed germline genetic data into the genetic ancestry principal components using weights derived for European, African, and Asian populations from the 1000 Genomes Project reference data (31). We further restricted our analysis to samples with < 10% inferred non-European ancestry.

### Identifying recurrently mutated cancer genes and hotspot mutations

We identified recurrently mutated cancer genes, defined as genes with ≥ 5% carriers of missense mutations, for each selected cancer type from the somatic data. Not all panel genes were sequenced on every sample (multiple panel versions exist); we thus further excluded those identified gene-cancer pairs with < 500 sequenced samples. We included additional genes that were identified as highly significantly mutated or significantly mutated genes among known cancer genes for each selected cancer type from the TumorPortal (http://www.tumorportal.org/) (32). A total of 135 cancer genes and 342 gene-cancer pairs were identified, with the mutation frequency ranging from 0.0036 to 0.73 (**Fig. 2A**; Supplementary Table S3). Mutation status for each sample and gene is defined as whether this sample carries at least one functional mutation (frame_shift_del, frame_shift_ins, frameshift, initiator_codon, missense and splice_region, missense_mutation, nonsense_mutation, protein_altering, splice_site, start_lost, stop_lost, and translation_start_site) in this gene and is considered to capture the “local” tumor mutation (mutation in one cancer gene).

**Figure 2.**
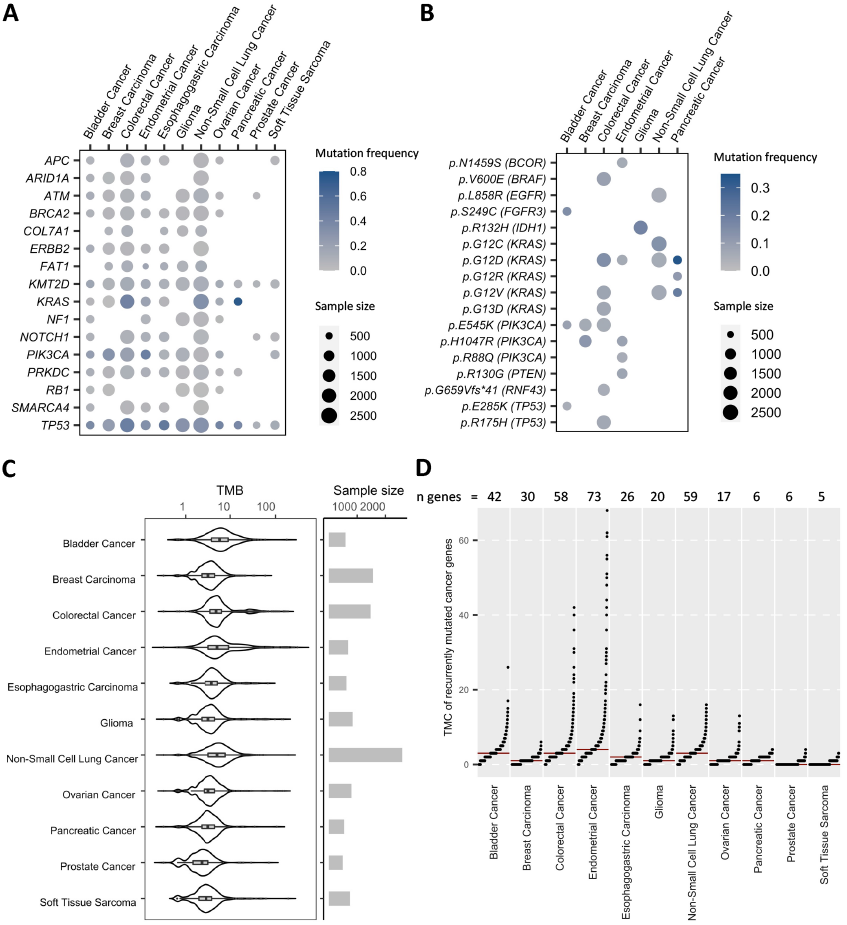
Local and global tumor mutations of 11 cancer types. **A**, Mutation frequency and sample size of the identified recurrently mutated cancer genes for each cancer type. A total of 135 genes were selected for 11 cancer types (Supplementary Table S3); only genes that are recurrently mutated in ≥ 5 cancer types are shown on this figure. **B**, Mutation frequency and sample size of the identified hotspot mutations for each cancer type. There are 17 hotspot mutations in 10 genes for 7 cancer types. **C**, Distribution of TMB of all panel genes across cancers with sample sizes. **D**, Distribution of TMC of recurrently mutated cancer genes across cancers. Each dot represents a sample. The red horizontal line represents the median of TMC for each cancer type. The total number of recurrently mutated cancer genes selected for each cancer is listed on the top of the figure.

For each selected cancer type, we further identified specific mutations with ≥ 5% carriers in the somatic data as hotspot mutations. Seven of the 11 cancer types harbor at least one hotspot mutation. A total of 17 hotspot mutations and 25 mutation-cancer pairs were identified, with the mutation frequency ranging from 0.051 to 0.33 (**Fig. 2B**; Supplementary Table S4). A binary variable of the local mutation status was created to indicate whether a sample carries a specific hotspot mutation.

### Quantifying TMB of all panel genes and TMC of recurrently mutated cancer genes

TMB is defined as the total number of missense mutations per megabase based on the targeted sequencing data of all panel genes (**Fig. 2C**). It captures the total mutations in all targeted cancer genes and is considered as a refined “global” mutational burden restricting to a set of cancer-related genes rather than the genome-wide mutational burden. In addition to TMB, we also calculated TMC for each sample, which is defined as the count of recurrently mutated cancer genes (specific to each cancer type) that harbor at least one missense mutation. The number of identified recurrently mutated cancer genes varies across cancer types (**Fig. 2D**). Compared to TMB, TMC is a more refined measure of the mutational burden in likely driver genes for a cancer. Moreover, by counting the genes instead of the mutations, the TMC analysis would be less sensitive to hypermutable outliers.

### Identifying eQTL from the Genotype-Tissue Expression (GTEx) Project for all selected genes

We obtained the eQTL and gene expression association results in normal tissue for all selected genes from the meta-analyzed multi-tissue eQTL results using METASOFT (33) from the GTEx Analysis V8 release. We selected those genome-wide significant eQTL with *P* < 5 × 10^−8^ from any of the fixed effect (FE), random effect (RE), or Han and Eskin’s random effect (RE2) models. Variants with minor allele frequency < 1% were further removed. A total of 28,486 eQTL for 114 genes with imputed germline data available were included in the analysis. We performed LD clumping with r^2^ = 0.3 on the final list of eQTL for each gene to identify independent loci, which was used to determine the number of effective tests in the association analyses (34).

### Assessing the associations of cancer gene eQTL with TMB and TMC

We assessed the association between each selected cancer gene eQTL and TMB of all panel genes for each cancer by fitting a linear model adjusting for age, gender (if applicable), panel version, and tumor purity. MSI status was also adjusted as a covariate for the models of colorectal and endometrial cancer where a substantial proportion of the cases display hypermutability (35,36). TMB was Winsorized to 98% within each cancer type to reduce the impact of potential outliers on the association results. The associations between cancer gene eQTL and TMC were evaluated for recurrently mutated cancer genes for each cancer type by fitting a negative binomial model with the same covariates as the TMB models. Sensitivity analysis was performed to assess the impacts of potential TMB or TMC outliers on the association results by varying the Winsorization thresholds and using standardized TMB. For TMC, we further evaluated the impacts of using count of missense mutations instead of count of mutated genes on the germline-somatic associations.

### Assessing the associations of cancer gene eQTL with recurrently mutated cancer genes and hotspot mutations

The local impact of each cancer gene eQTL on the risk of having somatic mutations in that gene or a nearby hotspot mutation was assessed using logistic regression. These analyses further adjusted for TMB along with all the covariates included in the TMB or TMC models. Meta-analysis was performed to evaluate the broad impact of a cancer gene eQTL on the mutation status of one gene or mutation across cancers.

## Data availability statement

The individual-level data used in this study are not publicly available due to patient privacy requirements. Other unidentifiable data generated in this study are available within the article and its supplementary data files.

## Results

### Germline cancer gene eQTL influence global tumor mutations

We analyzed 28,486 eQTL for 114 cancer genes and assessed their associations with TMB of all cancer genes sequenced on the panel across cancers. There were 1,317 independent eQTL (r^2^ < 0.3) after LD clumping. We identified 22 significant eQTL-TMB associations representing 3 independent gene-cancer pairs that passed the Bonferroni correction threshold accounting for the number of effective tests (*P* < 3.45 × 10^−6^; Supplementary Table S5). **Table 1** summarizes the results for the most significant association at each locus. There exists heterogeneity in the effects of these eQTL on TMB across cancers (Supplementary Table S6). Sensitivity analysis on the impacts of potential outliers showed that the association of the *GLI2* eQTL and TMB in ovarian cancer was sensitive to the changing Winsorization threshold (Supplementary Table S7). This association also became non-significant if we use standardized TMB as the outcome (beta = 0.26, *P* = 0.43) while the other two top associations remained nominally significant (beta = −2.33, *P* = 1.57 × 10^−3^ for rs139944315 (*WRN*) and TMB in glioma; beta = −0.23, *P* = 0.04 for rs11075646 (*CBFB*) and TMB in esophagogastric carcinoma).

**Table 1.**
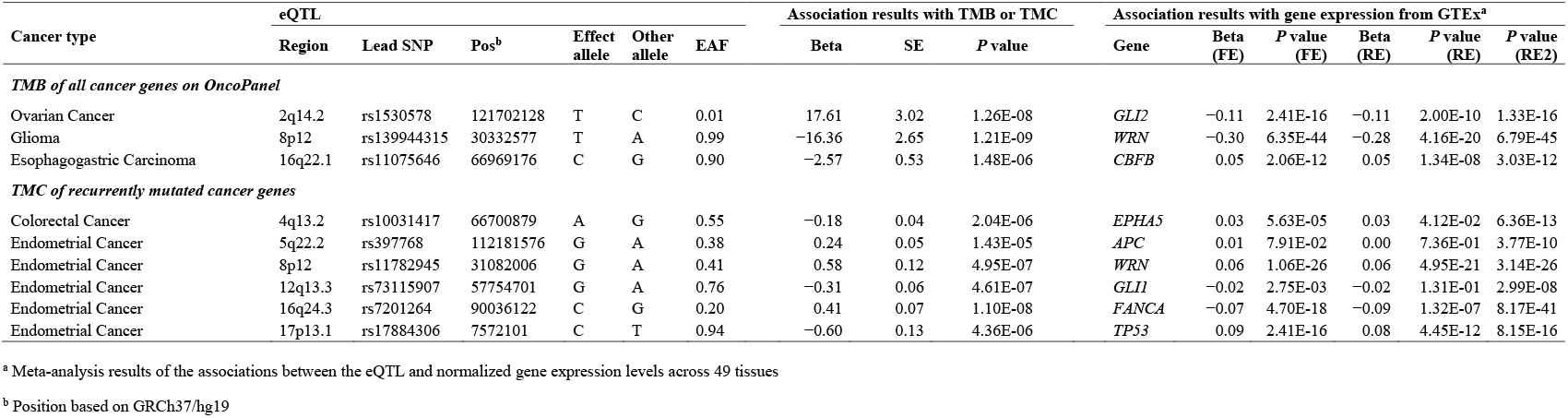
Significant associations between cancer gene eQTL and global tumor mutations.

To further investigate the relationship between the observed germline-somatic associations and gene expression, we compared our results with the association results between the identified top eQTL and expression level of the specific cancer genes in normal tissue in GTEx (**Table 1**; **Fig. 3**). The T allele of rs1530578 was associated with elevated TMB in ovarian cancer and reduced expression of *GLI2* across tissues (**Fig. 3A,D**). The largest effect of rs1530578 on *GLI2* expression was observed in ovary with beta = −0.55 and *P* = 3.93 × 10^−5^ (**Fig. 3A**). rs139944315 was associated with TMB in glioma and expression of *WRN* across tissues in a consistent direction (**Fig. 3B,E**). While the largest effect of this variant on *WRN* expression was observed in subcutaneous adipose tissue, there was also an association in putamen of basal ganglia with beta = −0.51 for the T allele and *P* = 0.05 (**Fig. 3B**). Finally, we found that the C allele of rs11075646 was associated with decreased TMB in esophagogastric carcinoma and slightly increased expression of *CBFB* across tissues (**Fig. 3C,F**). This variant had a nominally significant impact on *CBFB* expression in both gastroesophageal junction (beta = 0.10 and *P* = 0.02, **Fig. 3C**) and mucosa of esophagus (beta = 0.08 and *P* = 0.01) while the most significant effect was observed for thyroid (**Fig. 3F**). None of these three top variants or variants in high LD with them have been linked to cancer incidence in genome-wide association studies (GWAS) from GWAS Catalog (https://www.ebi.ac.uk/gwas/home) (37).

**Figure 3.**
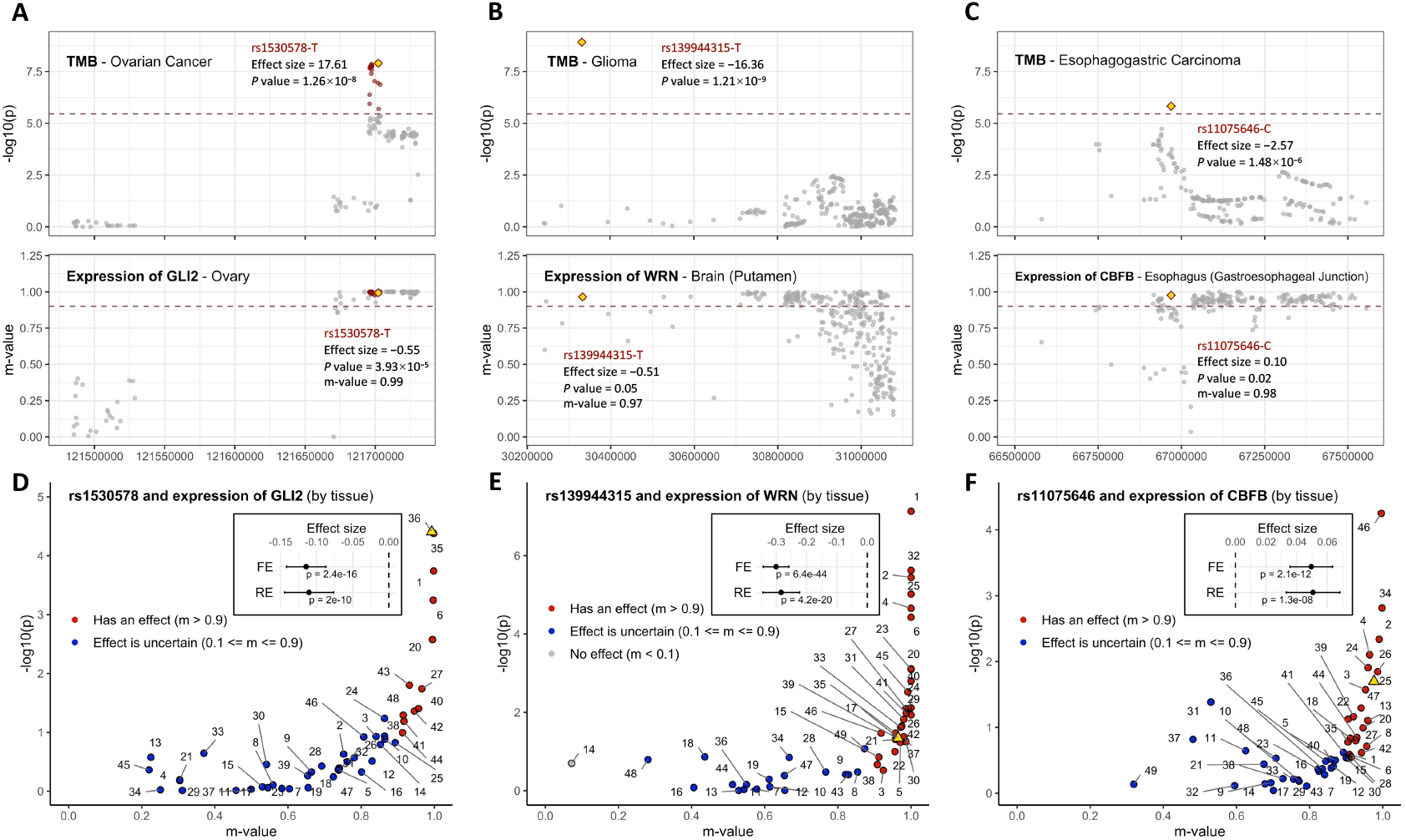
Associations of eQTL with TMB of all panel genes and gene expression across tissues. **A-C**, All selected eQTL for genes identified from the top eQTL-TMB associations are shown. Association results (−log_10_(*P*)) for eQTL and TMB are from linear models adjusting for age, gender (if applicable), tumor purity, and panel version. Association results (m-value, the posterior probability that an effect exists in a tissue) for eQTL and gene expression in the “matching tissue” are from GTEx; matching tissue was selected as the tissue with the largest m-value among all relevant tissues for the corresponding cancer type. Each dot represents a variant; variants that are significantly associated with both TMB and gene expression (in any meta-analysis model) are in red with the top variant marked as yellow diamond. RSID, effect allele, effect size, *P* value, and m-value for the top variant are annotated on the plots. The horizontal red dashed lines denote the significant threshold for TMB associations (*P* = 3.45 × 10^−6^) and “has an effect” threshold for gene expression associations in the matching tissue (m-value = 0.9). **D-F**, Association results of the top variants with expression level of the eQTL genes identified from the top eQTL-TMB associations by tissue from GTEx. The −log_10_(*P*) are from single-tissue eQTL analysis. Each dot represents a tissue with the matching tissue for the specific cancer marked as yellow triangle. Meta-analysis results across tissues from FE and RE models are provided on the plots. See Fig. 5 for tissue annotations.

We next assessed the impacts of cancer gene eQTL on TMC, which quantifies the mutational burden of a set of genes that are recurrently mutated in the specific cancer. There were 145 significant eQTL-TMC associations after Bonferroni correction (*P* < 1.73 × 10^−5^; Supplementary Table S8), representing six independent gene-cancer pairs (**Table 1**). Sensitivity analysis showed that all top TMC associations were robust to a wide range of Winsorization thresholds (Supplementary Table S9). Replacing count of mutated genes with count of mutations also yielded similar results compared to the main analysis (Supplementary Table S10). Given that all top eQTL-TMC associations were identified for colorectal and endometrial cancer, we further performed a stratified analysis by MSI status. There was no substantial deviation in the effect estimates for MSS or MSI subgroup from the main analysis though the subgroup results were less significant, especially for MSI samples, which was likely due to the reduced sample sizes (Supplementary Table S11). Finally, we compared these top eQTL-TMC associations to the previous eQTL-TMB results and found that all these top germline variants were associated with TMB in the corresponding cancers in consistent directions with TMC with nominal significance (Supplementary Table S12).

There exists substantial heterogeneity in the associations with gene expression level across tissues for many of the top variants in the TMC associations (**Fig. 4**). Two of the tissue-specific associations have both *P* < 0.05 and m-value > 0.8: rs10031417 and *EPHA5* expression in sigmoid colon and rs7201264 and *FANCA* expression in uterus (**Fig. 4A,E**). The A allele of rs10031417 was associated with lower somatic mutational burden in recurrently mutated cancer genes in colorectal cancer and slightly higher expression of *EPHA5* across tissues (**Fig. 4A,G**); this positive effect on *EPHA5* expression was larger in sigmoid colon with beta = 0.17 and *P* = 1.11 × 10^−3^ (**Fig. 4A**). It is worth noting that a variant that is in LD with rs10031417 (rs13104357, r^2^ = 0.18) has also been reported to be associated with *EPHA5* expression in colorectal tumor samples in The Cancer Genome Atlas (TCGA) (38); the direction of this association in tumor was consistent with in normal tissue. rs7201264-C allele was associated with both increased TMC in endometrial cancer and decreased *FANCA* expression across tissues (**Fig. 4E,K**); it had a specific significant impact on *FANCA* expression in uterus (**Fig. 4E**; beta = −0.28 for the C allele, *P* = 0.02). rs78378222, that is in LD with the top variant identified for TMC in endometrial cancer and *TP53* expression (rs17884306, r^2^ = 0.21 with rs78378222), has been previously associated with the risk of uterine fibroids and several cancers in but not the risk of endometrial cancer specifically (39,40).

**Figure 4.**
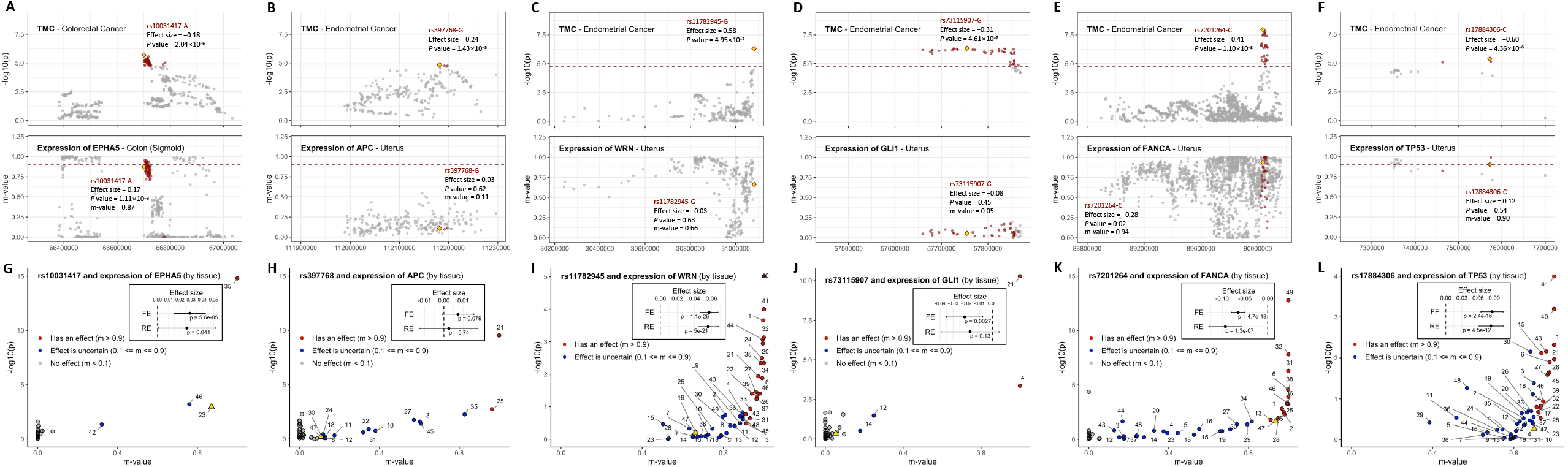
Associations of eQTL with TMC of recurrently mutated cancer genes and gene expression across tissues. **A-F**, All selected eQTL for genes identified from the top eQTL-TMC associations are shown. Association results (−log_10_(*P*)) for eQTL and TMC are from negative binomial models adjusting for age, gender (if applicable), tumor purity, panel version, and MSI status. Association results (m-value, the posterior probability that an effect exists in a tissue) for eQTL and gene expression in the “matching tissue” are from GTEx; matching tissue was selected as the tissue with the largest m-value among all relevant tissues for the corresponding cancer type. Each dot represents a variant; variants that are significantly associated with both TMC and gene expression (in any meta-analysis model) are in red with the top variant marked as yellow diamond. RSID, effect allele, effect size, *P* value, and m-value for the top variants are annotated on the plots. The horizontal red dashed lines denote the significant threshold for TMC associations (*P* = 1.73 × 10^−5^) and “has an effect” threshold for gene expression associations in the matching tissue (m-value = 0.9). **G-L**, Association results of the top variants with the expression level of the eQTL genes identified from the top eQTL-TMC associations by tissue from GTEx. The −log_10_(*P*) are from single-tissue eQTL analysis. Each dot represents a tissue with the matching tissue for the specific cancer marked as yellow triangle. Meta-analysis results across tissues from FE and RE models are provided on the plots. See Fig. 5 for tissue annotations.

### Local impacts of germline eQTL on somatic mutations in cancer genes

Investigation of the local impacts of eQTL for a cancer gene on somatic mutations in that gene is also of interest as it may point to a direct and testable mechanism of how germline variations modify the susceptibility to somatic events. None of the individual associations between somatic mutation status for recurrently mutated genes and their eQTL passed the Bonferroni correction threshold (*P* < 1.73 × 10^−5^). The most significant association observed was between a *TSC2* eQTL and somatic *TSC2* mutation status in endometrial cancer (beta = −1.81 for rs12918530-C allele, *P* = 1.56 × 10^−4^; Supplementary Table S13). Looking across all cancers, there was a significant (*P* < 6.91 × 10^−5^) association between an *ATM* eQTL (lead SNP: rs4753834 at 11q22.3) and somatic *ATM* mutations from a meta-analysis of 8 cancers (**Fig. 5**; Supplementary Table S14). The G allele of rs4753834 was associated with a lower risk of having somatic mutations in *ATM* (beta = −0.35, *P* = 3.43 × 10^−5^ across cancers from FE model) and increased expression of *ATM* in normal tissues (beta = 0.05, *P* = 1.03 × 10^−20^ across tissues from RE model). This variant also had specific effects on *ATM* expression in many tissues related the 8 cancers, including mammary tissue (beta = 0.06), sigmoid colon (beta = 0.09), hypothalamus (beta = 0.12), lung (beta = 0.07), and prostate (beta = 0.11), all with *P* < 0.05 and m-value > 0.9. Moreover, variants that are in LD with rs4753834 have also been associated with *ATM* expression in tumor samples of breast cancer (rs673281, r^2^ = 0.21, beta = −0.08 for the T allele, *P* = 1.98 × 10^−4^) and glioma (rs1003623, r^2^ = 0.21, beta = −0.11 for the T allele, *P* = 4.56 × 10^−4^) (38); the directions were also consistent with those in normal tissues. We additionally tested the associations of *ATM* eQTL and TMB or TMC of cancer genes and found that variants in LD with rs4753834 (lead SNP: rs672964, r^2^ = 0.21 with rs4753834) were associated with TMB (beta = −0.69 for rs672964-C, *P* = 2.97 × 10^−5^) and TMC (beta = −0.07 for rs672964-C, *P* = 0.02) in non-small cell lung cancer in the consistent direction with *ATM* mutation status. No association with cancer risk was found for rs4753834 or its tagging SNPs in GWAS Catalog.

**Figure 5.**
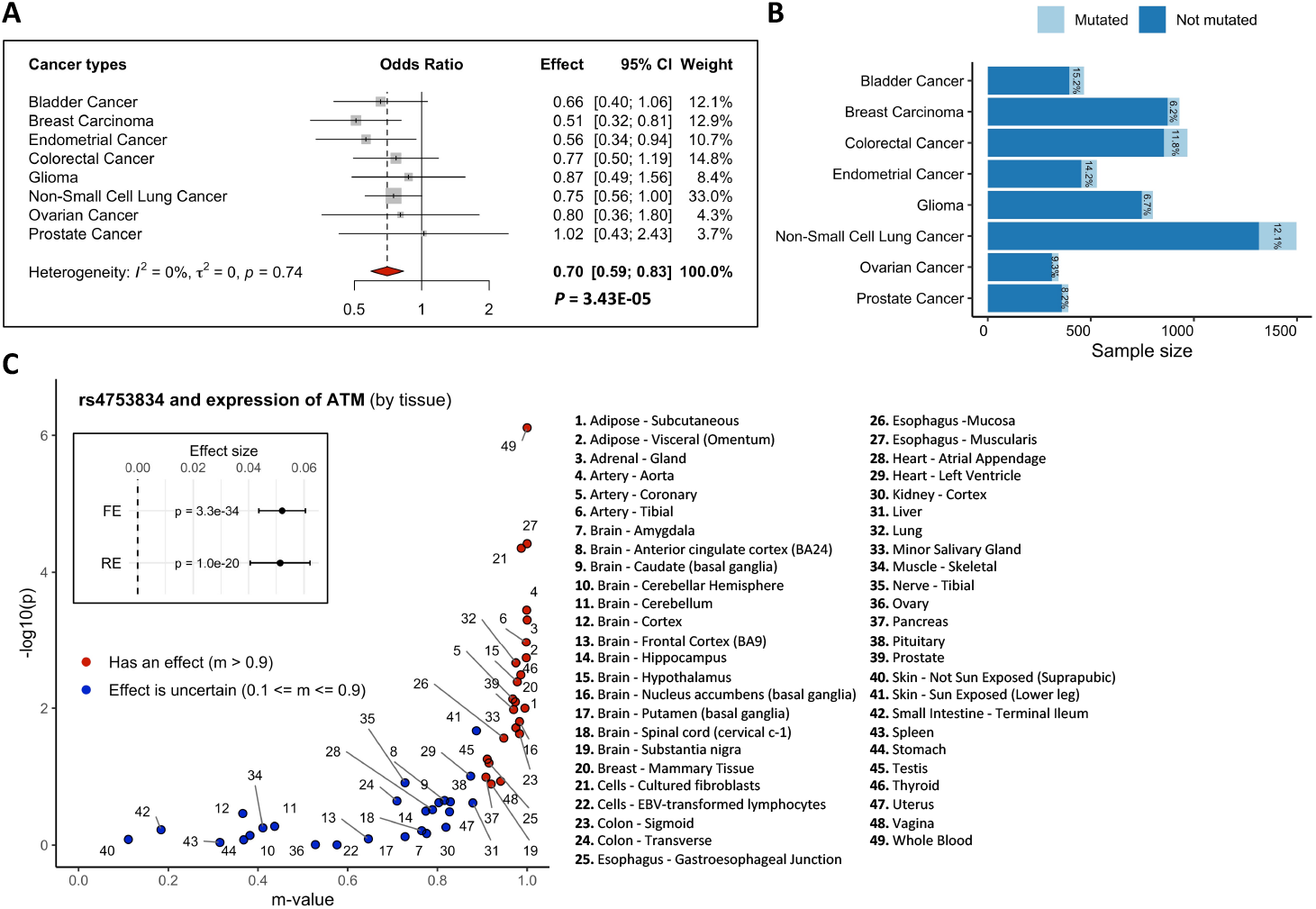
rs4753834 is associated with both *ATM* expression and somatic *ATM* mutations. **A**, Associations between rs4753834 and risk of having somatic mutations in *ATM* across 8 cancers. The odds ratio is associated with the G allele of rs4753834. Meta-analysis results from the fixed-effect model are shown. **B**, Sample sizes and mutation frequencies of the 8 cancer types. Note that these numbers are based on samples included in the final models. **C**, Association results of rs4753834 and *ATM* expression by tissue from GTEx. The −log_10_(*P*) are from single-tissue eQTL analysis. m-value is the posterior probability that an effect exists in a tissue. Results from the FE and RE meta-analysis across tissues are also shown on the plot.

We also identified nominal associations between eQTL for cancer genes identified in the global tumor mutation analysis with the somatic mutation status of that gene in the corresponding cancer. We found that rs1897693 (r^2^ = 0.42 with rs10031417) was associated with both the expression of *EPHA5* in normal tissues (beta = 0.03 for the C allele, *P* = 0.03 across tissues from RE model) and the somatic mutation status of *EPHA5* in colorectal cancer (beta = −0.66 for the C allele, *P* = 0.01). Another variant rs55671402 was associated with *FANCA* expression in normal tissues (beta = −0.13 for the C allele, *P* = 9.67 × 10^−13^ across tissues from RE model; beta = −0.54 for the C allele, m-value = 0.98, *P* = 1.35 × 10^−3^ in uterus) and somatic mutations in *FANCA* in endometrial tumors (beta = −1.23 for the C allele, *P* = 8.61 × 10^−3^).

We further assessed the impacts of eQTL for a cancer gene on each identified hotspot mutation in that gene. None of the associations passed the Bonferroni correction threshold (*P* < 3.40 × 10^−4^) with the most significant association observed for rs1867930 with p.S249C in *FGFR3* in bladder cancer (beta = 0.60 for the G allele, *P* = 3.54 × 10^−3^; Supplementary Table S15). Only one nominally significant (*P* < 0.05) association from the meta-analysis across cancers was found for rs11047823 with p.G12D in *KRAS* across colorectal cancer, endometrial cancer, non-small cell lung cancer, and pancreatic cancer (beta = 0.24 for the G allele, *P* = 0.01 across cancers from the FE model), though it still did not pass the Bonferroni correction threshold for significance (*P* < 5 × 10^−3^).

## Discussion

In this study, we systematically evaluated the influence of germline variants that are associated with cancer gene expression on somatic mutations in specific cancer genes across 11 cancer types, leveraging large-scale clinical targeted panel sequencing data, germline data imputed from tumor sequences, and cancer gene eQTL data from GTEx. Our analysis revealed novel associations of germline eQTL for well-established cancer genes with local mutation status of a single cancer gene or the global mutational burden. These findings provide the initial evidence for the hypothesis that germline variants can influence local and global tumor mutations by altering the expression level of specific cancer genes. The underlying molecular mechanisms of the identified associations can be further investigated through functional analysis and in cancer cell lines.

Although our findings are consistent with the putative mechanism that germline variants affect somatic mutations through gene expression, there are also other possible scenarios that can yield the same results (**Fig. 6**). First, given that there exists a causal impact of eQTL on somatic mutations, we still cannot conclude that this is only mediated by the transcript abundance of the specific eQTL gene. The germline eQTL may regulate the expression of other genes which contribute to somatic mutagenesis, or they might be associated with somatic mutations through other pathways that are not related to gene expression (**Fig. 6A**). Finding an eQTL signal in the cancer-related tissue can provide further support that gene expression plays a role in the germline-somatic relationship. Second, we are studying somatic mutations in developed tumor (S’) rather than in normal or precancerous tissue (S) (**Fig. 6A**). S’ can serve as a proxy for S, though it was measured after tumorigenesis and might be further influenced by other factors such as the tumor microenvironment (41). Here, we are studying mutations in cancer genes that have been identified as potential drivers for carcinogenesis. Even if some mutations in those genes occurred after cancer initiation, our results could still inform us of the role of germline variants in inducing somatic mutations during cancer progression. Finally, even when there is no direct causal effect of germline variants on somatic mutations, we may still observe this association among cancer patients. Consider the three possible scenarios in **Fig. 6B** given that a germline-somatic association was observed: germline variants may influence somatic mutation and they may or may not have an effect on cancer diagnosis through other pathways; however, under the situation that the germline variants only influence cancer diagnosis through other pathways and there is no causal effect on somatic mutations, we may still observe this germline-somatic association among cancer patients due to collider bias (**Fig. 6B**). We are unable to distinguish between these three scenarios based on our data, but we can leverage information from other sources (e.g., association results of the germline variants with cancer incidence from GWAS) to weigh these possible scenarios for each identified association.

**Figure 6.**
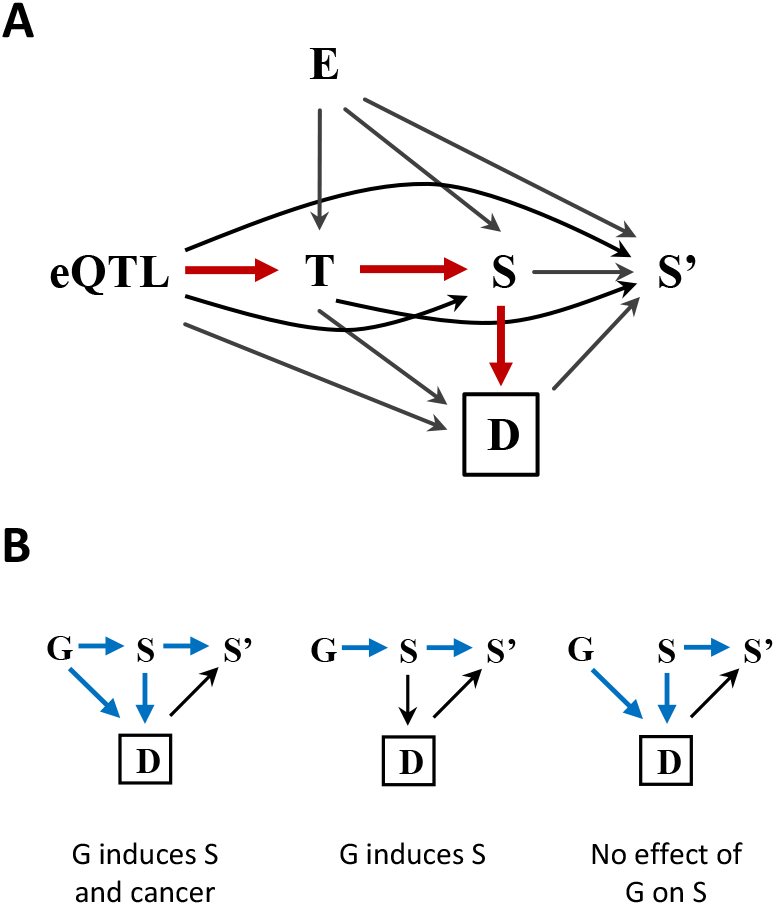
Hypothetical relationships between germline variants, cancer gene expression, somatic mutations, and cancer diagnosis. **A**, Complete relationships between germline eQTL, environmental factors (E), expression level of cancer genes in normal tissue (transcript abundance, T), somatic mutations in cancer genes in normal tissue before tumor development (S), cancer diagnosis (D), and somatic mutations in cancer genes in tumor after cancer diagnosis (S’). Our hypothesis is that germline eQTL regulate the expression of cancer genes; the transcript abundance of those cancer genes modifies the propensity of acquiring somatic mutations in those genes; having somatic mutations in those cancer genes is associated with an increased risk of cancer (the path shown by red arrows). Here, we are testing the associations between the eQTL and S’, which can serve as a proxy for S, among cancer patients (conditioning on D). **B**, Three possible relationships between germline variants (G), somatic mutations in normal tissue before tumor development (S), cancer diagnosis (D), and somatic mutations in tumor after cancer diagnosis (S’) given that an association between G and S’ is observed. Blue arrows on the graphs show the paths from G to S’ through S given that only cancer patients are included in the study (conditioning on D).

Most of the germline-somatic associations identified here were consistent with prior evidence, and many of them may be involved in the biological mechanisms that underlie patients’ response to immunotherapy. Among all the identified eQTL genes, *APC, ATM, CBFB*, and *TP53* have been predicted as pan-cancer tumor suppressor genes across 33 cancer types in TCGA (1). We observed that the germline variants associated with reduced expression of these tumor suppressor genes were associated with increased tumor mutations, except for *APC* where the eQTL association with gene expression was close to null across tissues (but still significant) with no effect in uterus (**Table 1**; **Fig. 3-5**). The *APC* gene encodes the adenomatous polyposis coli protein which plays an important role in the Wnt signaling pathway (42) and interacts with E-cadherin, which regulates cell adhesion (43). Mutations that inactivate APC lead to disruption of β-catenin degradation, resulting in its translocation into the nucleus and activation of the transcription of multiple genes, which triggers cancer development, including endometrial carcinogenesis (44). Active β-catenin signaling has been linked to resistance to anti-PD-L1/anti-CTLA-4 monoclonal antibody therapy in melanoma (45). A recent study found that germline pathogenic variants in *APC* are associated with elevated TMB (46). In our work, the minor allele of the lead SNP is also associated with higher TMC of recurrently mutated cancer genes, but the direction of its association with *APC* expression is not clear (**Table 1**; **Fig. 4**). Intuitively, we would assume a variant that downregulates the expression of a tumor suppressor gene to be associated with elevated risk of cancer and somatic mutational burden, but this assumption might be oversimplified as the oncogenic or tumor suppressive effect of a gene on carcinogenesis and on somatic mutational burden would depend on the signaling pathway that the gene involved in and may vary substantially across cancer types (47). Here, the major allele of rs397768 slightly downregulates *APC* expression across tissues, if this indicates activation of β-catenin signaling in endometrial carcinogenesis, then it should be associated with resistance to immunotherapy and reduced tumor mutations as we observed. However, this interpretation depends upon many variable components involved in this complex biological process; further study is needed to elucidate the molecular mechanisms underlying these associations.

*ATM* germline and somatic mutations have been linked to multiple cancers. Activated ATM protein kinase phosphorylates a few key proteins which activates DNA damage checkpoint, leading to its main tumor suppressive effect of cell-cycle arrest and apoptosis (48). A study of pathogenic germline variants in cancer identified two-hits events for *ATM* where a germline variant in *ATM* is coupled with a somatic mutation in the other copy of the gene in multiple cancers (49). They also found that *ATM* pathogenic variant carriers had lower *ATM* expression, which is in line with our finding that the minor allele of rs4753834 is associated with lower expression of *ATM* but higher risk of having somatic mutation in the gene (**Fig. 5**). Recent studies also reported that *ATM* mutations were associated with improved response to immune checkpoint blockade therapy (50,51). We observed this inverse relationship of *ATM* expression with both somatic *ATM* mutations across cancers and TMB in non-small cell lung cancer, which may support the potential role of *ATM* as a therapeutic target for promoting the response to cancer immunotherapy.

*TP53*, which encodes protein p53, is one of the most frequently mutated genes in cancer. Genetic alterations in the p53 stress response pathway can affect the tumor suppressive role of *TP53* and promote tumorigenesis (52). Results from a recent study demonstrated evidence for the interactive effects of a germline cancer risk variant, rs78378222, and somatic mutation status of *TP53* on cancer risk, prognosis, and drug responses (24). The C allele of rs78378222 has been linked to lower expression level of wild-type *TP53* in both normal tissue and tumor, which in turns reduce p53 cellular activity and lead to poorer overall survival of patients. In our analysis, we found that the minor allele of rs17884306, which is correlated with the C allele of rs78378222, was associated with higher TMB and lower *TP53* expression (**Table 1**; **Fig. 3**). One study highlighted the predictive value of somatic *TP53* mutations for benefit from anti–PD-1/PD-L1 immunotherapy in lung cancer (53). Our results may provide further insights into how inherited genetic predisposition can influence patients’ response to immunotherapy through its effect on *TP53* expression and somatic mutational burden.

Increased expression of *GLI2* in the hedgehog signaling pathway has been found to induce PD-L1 expression in gastric cancer and promote tumor resistance to immunotherapy (54). We identified a germline eQTL at 2q14.2 that upregulates *GLI2* and is associated with lower TMB in ovarian cancer; nominally significant associations were also found in esophagogastric carcinoma and glioma in the same directions (**Table 1**; **Fig. 3**; Supplementary Table S6). These findings may shed light on the underlying mechanism of the link between TMB and response to immunotherapy in these specific cancers.

Reduced *EPHA5* expression has been linked to lymph node metastasis, advanced TNM stage, and poor survival outcome in colorectal cancer, supporting its tumor suppressive role in this cancer (55). Recent work showed that having somatic *EPHA5* mutations is positively associated with TMB and response to immune checkpoint inhibitor therapy in lung cancer (56). We also identified consistent associations of an *EPHA5* eQTL at 4q13.2 with both somatic *EPHA5* mutations and the global tumor mutations in colorectal cancer. This eQTL influences *EPHA5* expression in colorectal cancer and normal colon tissue (**Table 1**; **Fig. 3**); the allele that was associated with reduced expression was also associated with increased somatic mutations.

Further studies are needed to characterize the potential interactive effect of these identified germline variants, *EPHA5* expression, and somatic *EPHA5* mutations in colorectal cancer.

Our study has several limitations. First, as mentioned above, we cannot easily distinguish between several possible scenarios of the causal relationships that may be consistent with the observed associations between germline eQTL and tumor mutations. We suggest future studies to further investigate these associations in normal tissue or precancerous lesions and incorporating haplotype-level information. Experimental validation is also necessary to confirm the putative mechanisms through gene expression for the identified associations. Second, the use of germline data imputed from off-target reads in tumor sequencing provides only a probabilistic estimate of the imputed variant. Although the validation analysis of imputed common germline variants against SNP array yielded high accuracy (26), it would still be important to validate these findings using direct germline genotyping. Third, our analysis focused on somatic mutations in the tumor, but we only included eQTL identified from normal tissue, which may miss tumor-specific eQTL effects (19). However, using normal tissue eQTL as the genetic instrument is more consistent with our hypothesis that eQTL alter gene expression in normal tissue contributes to somatic mutagenesis and tumor initiation. Where available, we also cross-referenced the eQTL results to those in corresponding tumor tissue and found the results had consistent direction with those in normal tissue. Finally, we only focused on missense and a few other functional mutations; future studies can further investigate the germline impact on somatic copy number alteration or structural variation through gene regulation.

In conclusion, we systematically investigated the impacts of germline cancer gene eQTL on somatic mutations in cancer genes across 11 cancer types. Our results indicate that germline variants that regulate the expression of cancer genes also influence local and global tumor mutations. These findings provide further evidence for the important role of gene expression regulation in germline-somatic associations and open avenues for future research on the molecular mechanisms underlying these associations that confer cancer risk and sensitize cancer to immunotherapy.

## Acknowledgements

This work was supported by National Cancer Institute grants R01CA227237 and R01CA244569 (to A. Gusev), and R01CA260352 (to P. Kraft), Phi Beta Psi Sorority, and Emerson Collective.

The Genotype-Tissue Expression (GTEx) Project was supported by the Common Fund of the Office of the Director of the National Institutes of Health, and by NCI, NHGRI, NHLBI, NIDA, NIMH, and NINDS. The data used for the analyses described in this manuscript were obtained from the GTEx Portal on 08/04/2021.

